# The MorbidGenes panel: a monthly updated list of diagnostically relevant rare disease genes derived from diverse sources

**DOI:** 10.1101/2024.04.15.24305833

**Authors:** Robin-Tobias Jauss, Bernt Popp, Joachim Bachmann, Rami Abou Jamra, Konrad Platzer

**Affiliations:** Institute of Human Genetics, University of Leipzig Medical Center, 04103 Leipzig, Germany; Berlin Institute of Health at Charité, Universitätsmedizin Berlin, Center of Functional Genomics, 10115 Berlin, Germany

**Keywords:** virtual panel, exome, rare disease genes, routine diagnostics

## Abstract

**Purpose:** With exome sequencing now standard, diagnostic labs are in need of a, in principle, to-the-day-accurate list of genes associated with rare diseases. Manual curation efforts are slow and often disease specific, while efforts relying on single sources are too inaccurate and may result in false-positive or false-negative genes.

**Methods:** We established the MorbidGenes panel based on a list of publicly available databases: OMIM, PanelApp, SysNDD, ClinVar, HGMD and GenCC. A simple logic allows inclusion of genes that are supported by at least one of these sources, providing a list of all genes with diagnostic relevance.

**Results:** The panel is freely available at https://morbidgenes.uni-leipzig.de and currently includes 4960 genes (as of March 2024) with minimally sufficient evidence on disease causality to classify them as diagnostically relevant.

**Conclusion:** The MorbidGenes panel is an open and comprehensive overview of diagnostically relevant rare disease genes based on a diverse set of resources. The panel is updated monthly to keep up with the ever increasing number of rare disease genes.

## Introduction

Accurate and early genetic diagnosis can improve individual outcomes and lower costs for the health care system^1^. With exome sequencing now standard in many labs in rare disease diagnostics, there is an urgent need for an up-to-date accurate list of diagnostically relevant genes to evaluate. However, disease specific virtual panels overlaid on exome data might lead to false-negative results due to the omission of relevant genes, as such panels are time consuming to keep up to date. Similarly, genes with heterogeneous disease associations need to be considered for multiple panels, increasing the curation workload. Such a curation effort can practically only be done by larger labs or crowdsourced platforms like PanelApp^2^, as opposed to smaller labs in routine diagnostics. In our experience, it became increasingly evident that using small curated panels can be feasible in diagnostic workflows for specific phenotypes, but one single broad gene panel that included every gene with minimally sufficient evidence on disease causality also needs to be included in exome analyses, especially for heterogeneous diseases.

Comprehensive lists of diagnostically relevant genes do exist, e.g. the Mendeliome panel from PanelApp Australia. Also, efforts to harmonize gene-disease associations across diagnostic labs have launched recently^3^, but do lag behind in curation of disease genes. In addition, the differences between labs are likely stark as reflected by a recent study comparing exome reports across 41 labs^4^. A comprehensive framework for broader gene selection of virtual panels has been published recently^5^, but lacks continuous updates and implies a potential risk of missing variants in genes with evidence yet to come. This motivated us to establish an automated and open list of diagnostically relevant genes generated by using diverse sources that we called the MorbidGenes Panel.

## Materials and methods

To include all genes with minimally sufficient evidence on their disease causality, we collect data on a monthly basis from the publicly available and widespread databases OMIM^6^, ClinVar^7^, HGMD^8^, PanelApp^2^ and GenCC^3^. As neurodevelopmental disorders are a very frequent referral reason for genetic diagnostics, we decided to include SysNDD as an expertly curated phenotype specific database as well^9^. The genes are scored based on the available evidence and are included in the panel if at least one of the following criteria is fulfilled (Figure 1): 1. The gene has an OMIM disease (excluding provisional associations and susceptibility phenotypes), 2. ClinVar lists at least four (likely) pathogenic variants excluding copy number variants, 3. HGMD lists at least four pathogenic variants, 4. A green curation status in at least one panel either from the Genomics England PanelApp or the PanelApp Australia, 5. A status of “definite” in the SysNDD database or 6. at least one entry with status “definite” in GenCC. The MorbidGenes panel is updated on a monthly basis with a custom R package (see Data and code availability). A download interface allows users to access and filter the panel and incorporate the genes into their in-house pipelines.

**Figure 1:**
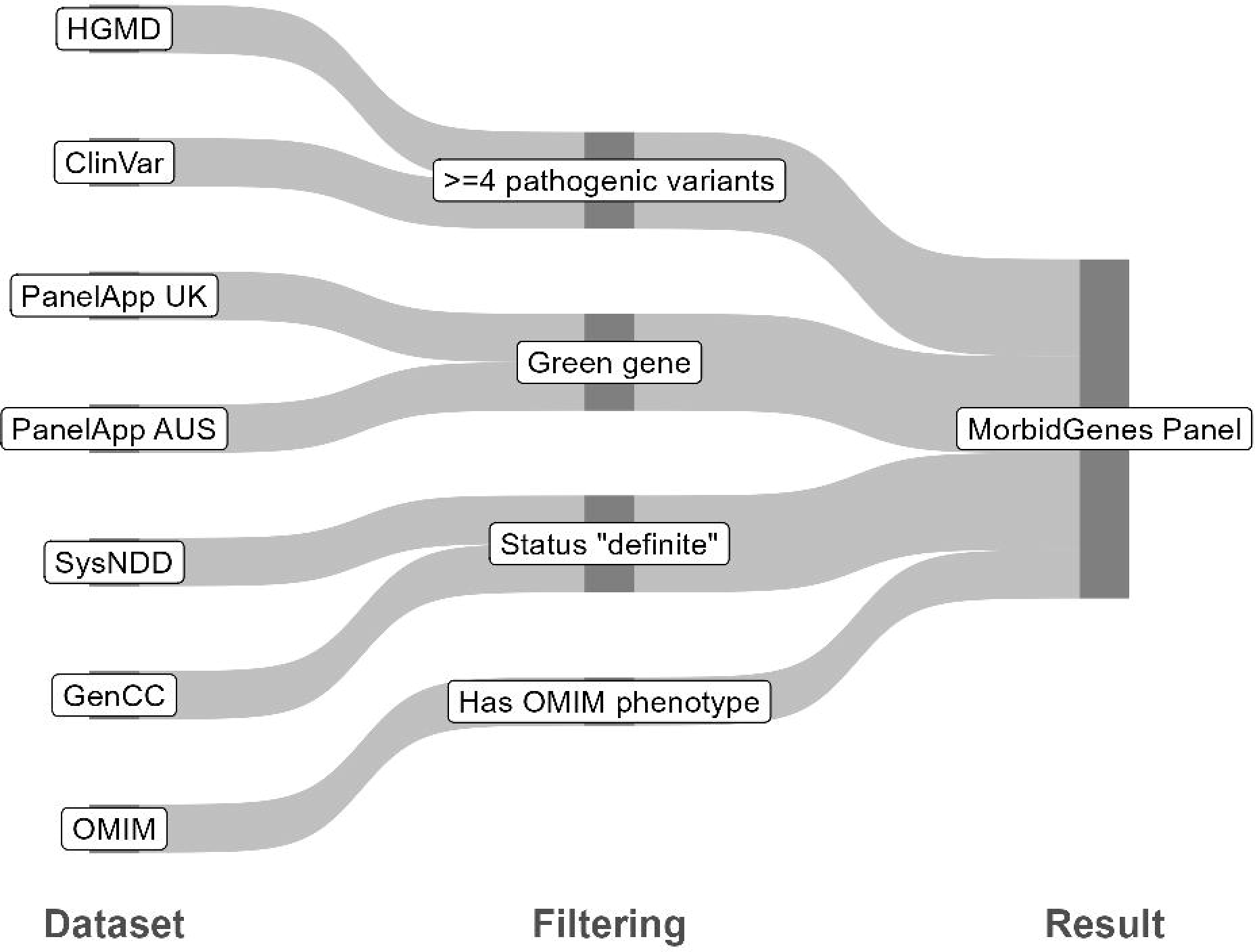
Flowchart of incorporated datasets and their respective filtering step. A gene is considered to be a Morbid Gene and included in the panel if it passes at least one of the filtering steps.

## Results

The MorbidGenes Panel is freely available at https://morbidgenes.uni-leipzig.de (Supplementary Figure 1) and currently contains 4960 genes (as of March 2024). On average, about 12 genes are added per month (Figure 2A). Each database awards a single point to the Morbidscore of a gene. Most of the gene-disease associations are well supported by multiple databases, reflected by the Morbidscore (Figure 2B). The largest group of genes comes with sufficient evidence on disease causality based on all sources ClinVar, HGMD, OMIM, GenCC and PanelApp without SysNDD (Figure 2C). Two thirds of all genes in the panel are supported by at least four databases, i.e., a Morbidscore of 4.

**Figure 2:**
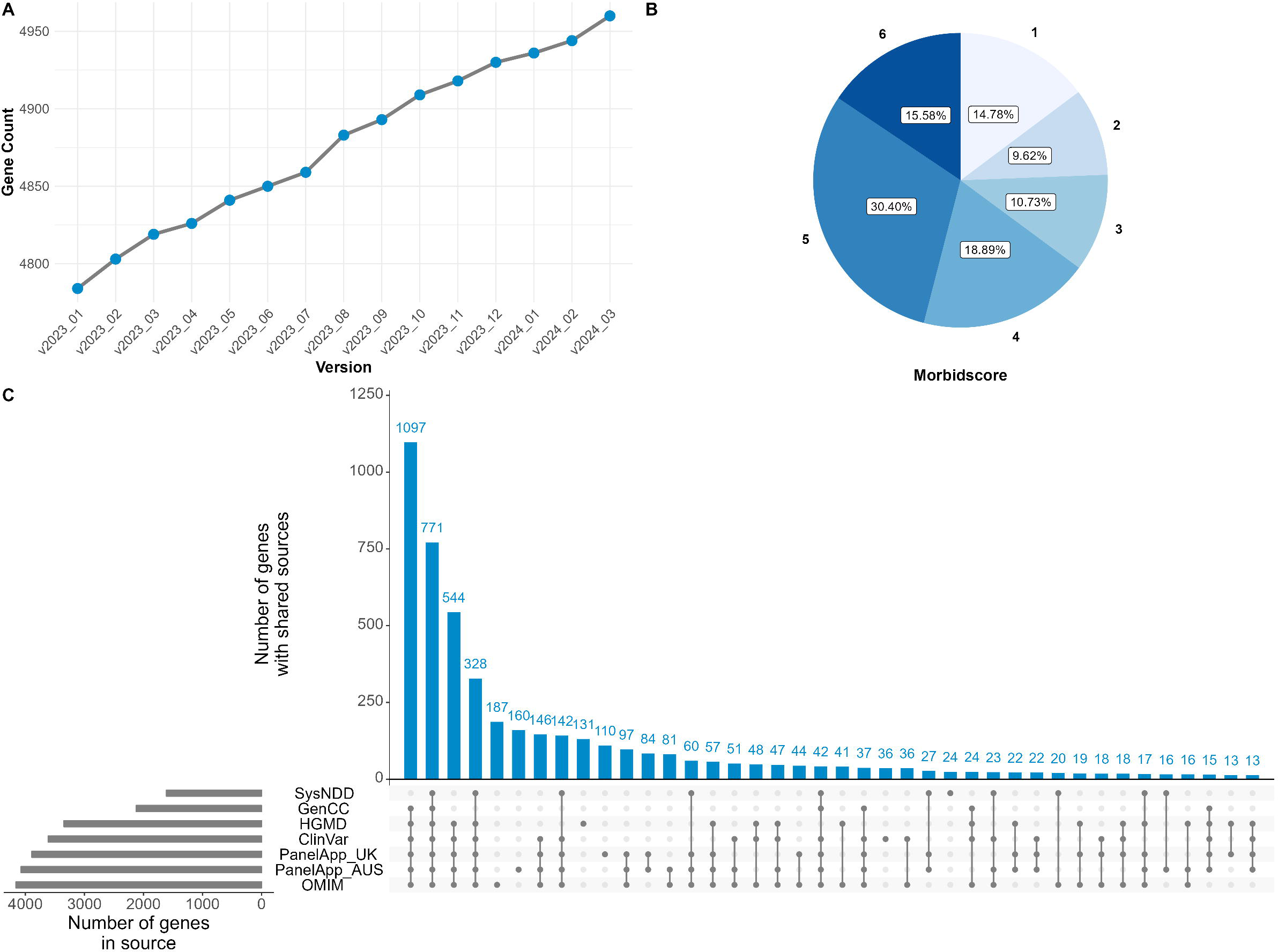
(A) Number of genes per MorbidGenes panel version. More than 130 genes were added to the panel from January to December 2023. (B) Distribution of the Morbidscore (in v2024_03). About two thirds of the genes contained in the MorbidGenes panel have a score of at least four, meaning that their disease association is supported in at least four of the databases included in the panel (C) UpSet plot of v2024_03 providing the number of genes per source (grey bars) and the number of genes sharing sources (blue bars and intersection matrix). Most genes are considered with sufficient evidence in ClinVar, HGMD, OMIM and PanelApp with or without SysNDD.

In total, 733 genes are supported by only one database. Of these, 187 are only supported by an OMIM entry. Some of these OMIM entries are based on a single, but recent publication (e.g. *VPS50* or *SLC30A7*), but no or limited pathogenic variants have been submitted to ClinVar or the gene has not been reviewed yet by platforms like PanelApp. On the other hand, the OMIM entries of some genes like *DIABLO* and *TNC* are based on a single publication from more than ten years ago, which describe a phenotype from a single family and have been marked with limited evidence by curation platforms like PanelApp and GenCC.

More than 340 genes have a green curation status in PanelApp, but no associated OMIM phenotype nor more than four pathogenic variants in HGMD or ClinVar. The Genomics England PanelApp and the PanelApp Australia share 3610 green genes, while 282 and 437 morbid genes are exclusively present in the Genomics England PanelApp and the PanelApp Australia, respectively. For some of these genes the evidence awarded to apply a green status it is not traceable.

## Discussion

The MorbidGenes Panel provides a comprehensive dataset of disease associated genes, aiding diagnostic labs with a monthly updated list of genes relevant for routine diagnostics.

Lists of diagnostically relevant genes manually curated by dedicated reviewers, like the Mendeliome panel by PanelApp Australia with currently more than 3800 green genes are in fact needed, but the huge efforts undertaken to maintain such a list are reflected by the nearly daily activity of the 65 reviewers of the panel. In addition, global efforts to share gene-disease lists by the global community like GenCC are highly commendable, but lag behind in curation and thus rather represent a 2^nd^ level of evidence. As GenCC gathers evidence from different data sources but does not generate new evidence itself, the GenCC database is in fact not needed for the curation of the MorbidGenes panel because it does not add unique genes to the panel. Still, the displayed evidence in GenCC provides a comprehensive overview of all available gene-disease curations, and for this reason the link to the available GenCC entry was retained in the MorbidGenes panel.

The drawback of our approach here is the loss of resolution, as only the gene name and the respective sources are retained. This is in contrast to manually curated panels like PanelApp, which provide definite gene-disease associations, clinical synopses, mode of inheritance and links to publications supporting this evidence. However, the MorbidGenes Panel does not aim to be another curated gold-standard, but rather serves as a first-in-line tool for a fast detection of clinically relevant genes that need to be integrated into genetic diagnostic routine. The drawback is compensated by providing links to the respective sources on the website.

Applying the MorbidGenes Panel as an *in silico* panel in routine diagnostics reduces the number of variants to evaluate from about 60.000 to about 20.000 per exome, eliminating variants in diagnostically irrelevant genes. We are aware of the fact that our MorbidGenes Panel still may include false positive genes, e.g. based on old and unconfirmed OMIM entries. As we wanted to establish a simple logic for inclusion of a gene and also keep manual curation to a minimum, we decided to also keep genes in the MorbidGenes panel with a Morbidscore of 1 only. A gene may only be supported by limited evidence based on a single publication with, for example, five individuals with a *de novo* missense variant – but if the analyzed individual has one of the exact same variants as reported in said publication and an overlapping phenotype, this would trigger an inclusion of the variant in the genetic report. Although evidence is limited on a number of genes, a case specific evaluation of the genotype and phenotype of the individual is always needed to decide which variants are to be reported back to the referring clinicians. As the Morbidscore represents the number of databases that support the evidence for a certain gene, the user can set an individual threshold to filter more stringently for genes with a minimal number of supporting databases.

As Figure 2 illustrates, it is of particular importance to include a heterogeneous set of data sources in panel curation, as important genes can be missed when focusing only on evidence from single databases. This is especially true for manually curated panels, as 282 and 437 genes are exclusively curated as a green gene in the Genomics England PanelApp and the PanelApp Australia, respectively, further undermining the need of diverse datasets. Regular updates are another important pillar in panel management, as new disease causing genes emerge continuously. More than 130 morbid genes were added from January to December 2023 (Figure 2A), each representing a potential diagnosis which might be missed if panels are not regularly updated. It is therefore crucial in routine diagnostics not only to include genes with minimal sufficient evidence, but also to keep the diagnostic pipelines up-to-date. Both aspects can be easily realised with our MorbidGenes Panel.

Our framework provides a broad list of diagnostically relevant genes based on a simple logic that limits curation workload and offers reproducible data generation which allows updates with even shorter time frames if necessary. The panel has been successfully implemented into the routine diagnostics at the Institute of Human Genetics of the University of Leipzig Medical Center with more than 2200 diagnostic exome reports a year. We are encouraging the genetics community to submit evidence on gene-disease associations in public databases as soon as possible, increasing the number of clinically relevant genes in automated virtual panels like the MorbidGenes panel.

## Data Availability

The panel is freely available at https://morbidgenes.uni-leipzig.de/. Code for regular panel updates and creation of gene lists is available at https://github.com/HUGLeipzig/MorbidUpdateR.

https://github.com/HUGLeipzig/MorbidUpdateR

## Supplemental information

*Supplementary Figure 1: Screenshot of https://morbidgenes.uni-leipzig.de. The web interface provides a dropdown menu to filter for specific panel versions and a download button for the displayed panel. The grid interface contains links to the respective data sources*.

## Acknowledgements

We thank the entire staff of the Institute of Human Genetics of the University of Leipzig Medical Center for their tireless work on rare disease diagnostics, counselling and research and for their input in improving the MorbidGenes Panel

## Author contributions

Conceptualisation: B.P., R.A.J.; Data curation: R.-T.J, B.P.; Formal analysis: R.-T.J., B.P.; Methodology: R.-T.J., B.P., J.B.; Software: R.-T.J., B.P., J.B.; Visualisation: R.-T.J.; Supervision: R.A.J., K.P.; Writing – original draft: R.-T.J., K.P.; Writing – review and editing: B.P., J.B., R.A.J. All authors reviewed and approved the final manuscript.

