## Supplementary figures and images for "The MorbidGenes panel: a monthly updated list of diagnostically relevant rare disease genes derived from diverse sources"

### Supplementary Figure 1

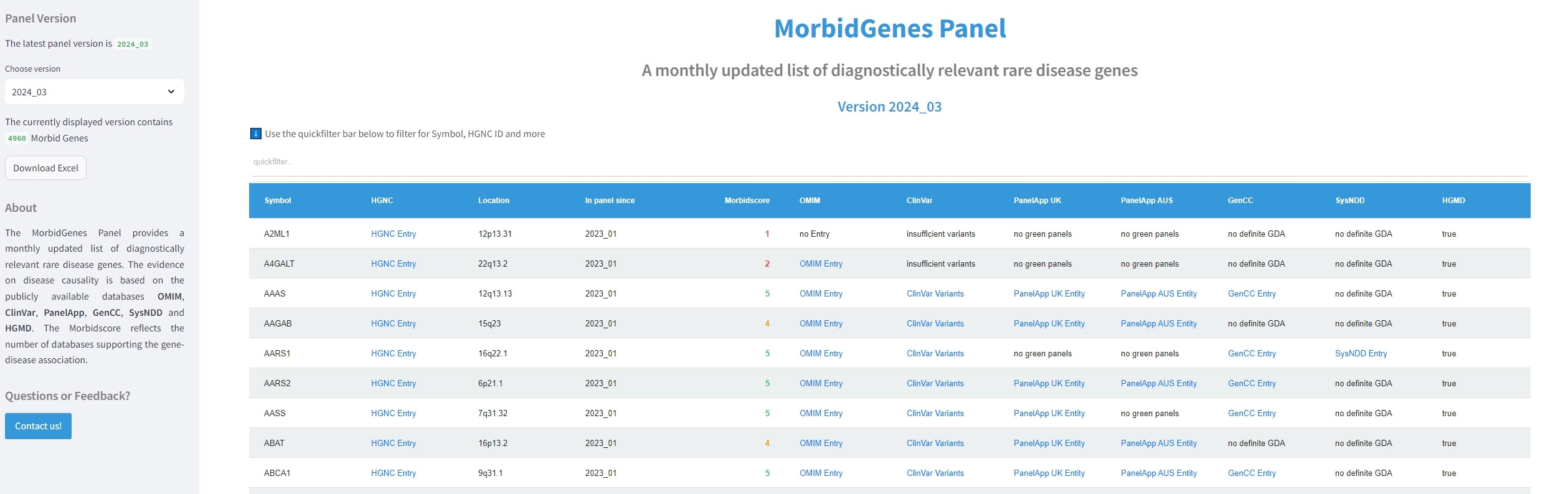
